# Diagnostic Performance of ChatGPT to Perform Emergency Department Triage: A Systematic Review and Meta-analysis

**DOI:** 10.1101/2024.05.20.24307543

**Authors:** Navid Kaboudi, Saeedeh Firouzbakht, Mohammad Shahir Eftekhar, Fatemeh Fayazbakhsh, Niloufar Joharivarnoosfaderani, Salar Ghaderi, Mohammadreza Dehdashti, Yasmin Mohtasham Kia, Maryam Afshari, Maryam Vasaghi-Gharamaleki, Leila Haghani, Zahra Moradzadeh, Fattaneh Khalaj, Zahra Mohammadi, Zahra Hasanabadi, Ramin Shahidi

## Abstract

**Background:** Artificial intelligence (AI), particularly ChatGPT developed by OpenAI, has shown potential in improving diagnostic accuracy and efficiency in emergency department (ED) triage. This study aims to evaluate the diagnostic performance and safety of ChatGPT in prioritizing patients based on urgency in ED settings.

**Methods:** A systematic review and meta-analysis were conducted following PRISMA guidelines. Comprehensive literature searches were performed in Scopus, Web of Science, PubMed, and Embase. Studies evaluating ChatGPT’s diagnostic performance in ED triage were included. Quality assessment was conducted using the QUADAS-2 tool. Pooled accuracy estimates were calculated using a random-effects model, and heterogeneity was assessed with the I² statistic.

**Results:** Fourteen studies with a total of 1,412 patients or scenarios were included. ChatGPT 4.0 demonstrated a pooled accuracy of 0.86 (95% CI: 0.64-0.98) with substantial heterogeneity (I² = 93%). ChatGPT 3.5 showed a pooled accuracy of 0.63 (95% CI: 0.43-0.81) with significant heterogeneity (I² = 84%). Funnel plots indicated potential publication bias, particularly for ChatGPT 3.5. Quality assessments revealed varying levels of risk of bias and applicability concerns.

**Conclusion:** ChatGPT, especially version 4.0, shows promise in improving ED triage accuracy. However, significant variability and potential biases highlight the need for further evaluation and enhancement.

## Introduction

Artificial intelligence (AI) has rapidly become a cornerstone of modern healthcare, revolutionizing various aspects of medical practice and research. AI technologies, including machine learning and natural language processing, have significantly improved diagnostic accuracy, treatment planning, and patient management. For instance, AI-driven systems can analyze vast amounts of medical data to identify patterns that are often undetectable to human clinicians, leading to earlier and more accurate diagnoses (1).

ChatGPT, developed by OpenAI, represents a significant milestone in the evolution of conversational artificial intelligence. Initially released in November 2022 and powered by the GPT-3.5 architecture, ChatGPT quickly demonstrated its capabilities in generating coherent and contextually relevant responses across various domains. The model was trained on extensive datasets, enabling it to perform tasks such as code generation, text summarization, and complex problem-solving with remarkable accuracy (2, 3). Also, ChatGPT can assist in generating medical documentation, providing accurate and efficient patient information, and even aiding in diagnostic processes by analyzing patient data and suggesting possible diagnoses. Moreover, ChatGPT’s ability to handle large volumes of data quickly and accurately makes it a valuable tool for streamlining workflows and improving overall healthcare efficiency (4).

Triage plays a crucial role in emergency departments (EDs) by prioritizing patients based on the severity of their conditions, ensuring that those who need urgent care receive it promptly. This process is essential for managing the high volume of patients and maintaining efficient workflow in busy EDs. The implementation of advanced triage protocols has been shown to improve patient outcomes and reduce waiting times, enhancing the overall efficiency of emergency care. Additionally, effective triage systems can help in better resource allocation, ensuring that critical cases receive immediate attention while non-urgent cases are managed appropriately (5).

The COVID-19 pandemic has highlighted some limitations of current triage systems, particularly in handling public health emergencies. The surge in patient numbers during the pandemic exposed weaknesses in existing triage protocols, including inadequate resources and the need for rapid adaptability. Traditional triage systems were not designed to cope with such unprecedented demand, leading to increased patient wait times and strained healthcare facilities. Moreover, there were ethical and logistical challenges in triage decisions, as protocols often failed to address disparities and ensure equitable care for all patients, particularly those from vulnerable populations (6, 7).

However, Evaluating the diagnostic performance of ChatGPT and it’s safety for emergency department (ED) triage is essential due to its potential to significantly enhance clinical decision-making and patient management. Studies have shown that while ChatGPT can achieve impressive diagnostic accuracy, it still faces challenges such as high rates of unsafe triage decisions compared to other systems like Ada and WebMD Symptom Checkers. Ensuring the accuracy and safety of ChatGPT in triage can help prevent misdiagnosis and improve patient outcomes, making it a valuable tool in emergency medicine (8).

Thus, this study aims to assess the diagnostic performance of ChatGPT and it’s safety in performing emergency department triage. By evaluating the accuracy, reliability, and effectiveness of ChatGPT in prioritizing patients based on urgency, this study seeks to provide a comprehensive understanding of its potential role in emergency care settings.

## Methods

Our study was conducted following the PRISMA (Preferred Reporting Items for Systematic Reviews and Meta-Analyses) guidelines (9). The protocol for this systematic review and meta-analysis was registered in the PROSPERO database with the registration code (CRD42024531858).

## Search Strategy

A comprehensive literature search was conducted across four major databases: Scopus, Web of Science, PubMed, and Embase. The search strategy was designed to identify studies evaluating the diagnostic performance of ChatGPT in emergency department (ED) triage. Keywords included combinations of terms such as “ChatGPT,” and “triage”. The search was restricted to studies published from the inception of these databases until 19 March 2024. Reference lists of the identified studies and relevant reviews were also screened to ensure completeness.

## Selection Process and Inclusion Criteria

The selection process involved two independent reviewers who screened titles and abstracts of all identified records. Disagreements were resolved through discussion, and a third reviewer was consulted if necessary. Full-text articles were retrieved for records deemed potentially relevant. Studies were included if they Evaluated the diagnostic performance of ChatGPT in emergency department triage. Studies were excluded if they were letters to the editor, guidelines, reviews, or did not focus on triage performance.

## Data Extraction Process

Two reviewers independently extracted data, including the first author, publication year, study design, type of medical cases triaged, number of evaluated scenarios or patients, country, version of used ChatGPT, comparison group, ChatGPT’s diagnostic performance, inter-rater reliability, main findings, and study limitations. Data were extracted using a predefined Excel format document. Discrepancies were resolved through discussion or by consulting a third reviewer.

## Quality Assessment

The quality of included studies was assessed using the QUADAS-2 (Quality Assessment of Diagnostic Accuracy Studies) tool (10). This tool evaluates the risk of bias in four domains: patient selection, index test, reference standard, and flow and timing. Applicability concerns were also assessed for each domain except flow and timing. Two reviewers independently performed the quality assessment, and disagreements were resolved through discussion.

## Statistical Analysis and Data Synthesis

Statistical analyses were conducted using R version 4.4. The diagnostic performance of ChatGPT was evaluated using the metaprop function to generate a pooled estimate of accuracy for triaging patients. a random-effects model was employed to account for variability among studies. Heterogeneity was assessed using the I² statistic, with values above 50% indicating substantial heterogeneity. Publication bias was evaluated using funnel plots. Sensitivity analyses were conducted to assess the robustness of the pooled estimates by excluding studies with a high Heterogeneities.

## Results

### Literature Review

A comprehensive literature search identified a total of 162 records across four databases: Scopus, Web of Science, PubMed, and Embase. After removing 92 duplicates, 70 records remained for screening. Through title and abstract screening, 27 records were excluded, leaving 43 full-text articles for detailed assessment. Of these, 29 were excluded for various reasons, such as being letters to the editor, guidelines, or not relevant to triage. Ultimately, 14 studies were included in our systematic review and meta-analysis **(Figure 1).**

**Figure 1.**
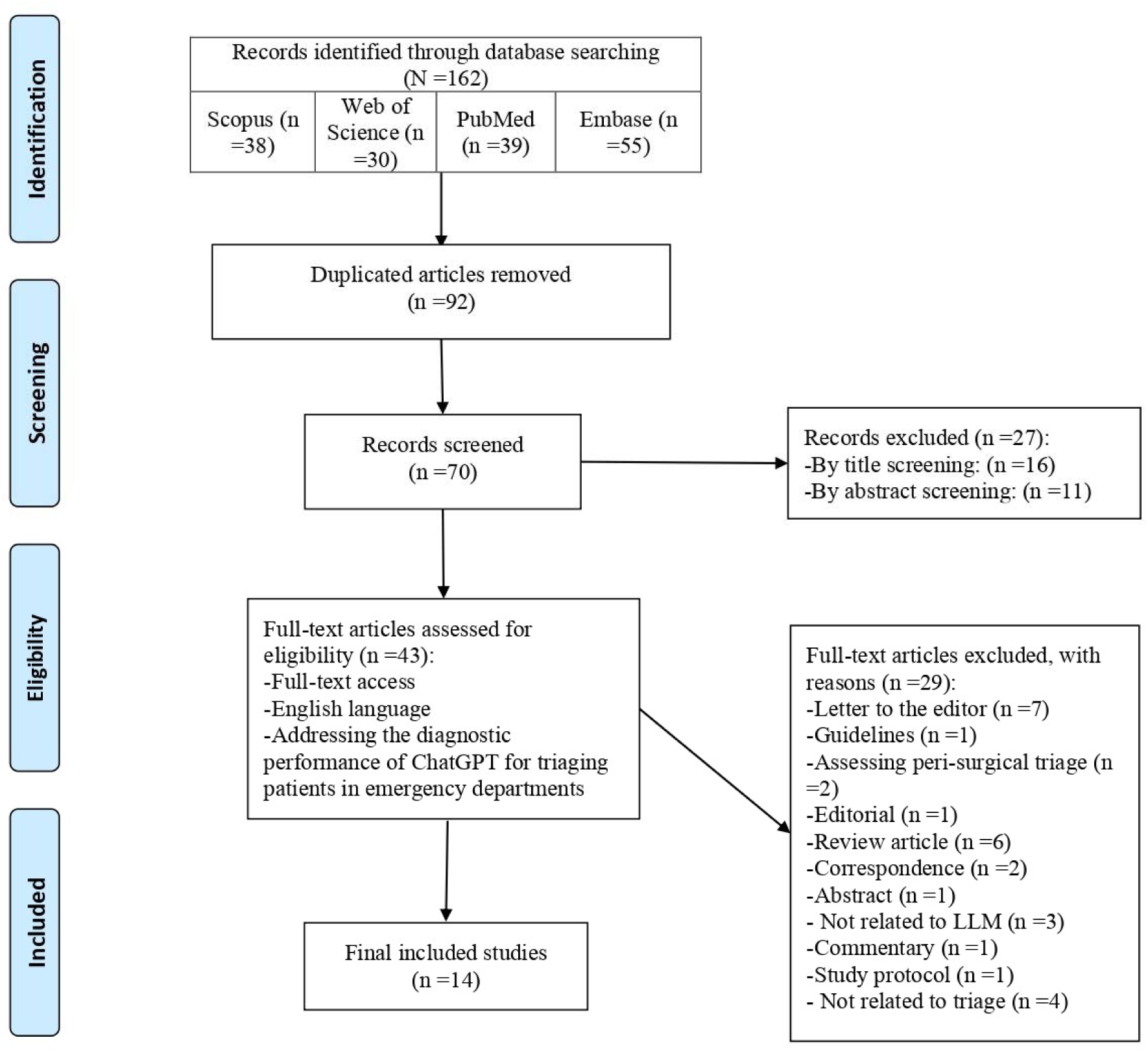
PRISMA Flowchart.

### Study Characteristics

Across these 14 included studies, a total of 1,412 patients or scenarios were evaluated. These studies were conducted between 2023 and 2024. The included studies predominantly employed a cross-sectional study design, focusing on a variety of medical cases in emergency departments (EDs). Specifically, seven studies addressed common cases in EDs (8, 11–16), two studies focused on mass casualty incidents (17, 18), one study examined cases needing neurosurgical attention (19), one study evaluated pre-hospital basic life support and pediatric advanced life support cases (20), two studies dealt with ophthalmic conditions (21, 22), and one study focused on metastatic prostate cancer patients (23).

Regarding the large language models used, six studies exclusively utilized ChatGPT-3.5 (12, 14–18), four studies used ChatGPT 4.0 (11, 21–23), and four studies employed both versions of ChatGPT in their analyses (8, 13, 19, 20). Inter-rater reliability varied among the studies. One study reported near-perfect agreement with the gold standard, with a Cohen’s kappa of 0.899 (11). Another study noted an agreement in MTS code assignment between the comparison group and ChatGPT-3.5, with a Cohen’s kappa of 0.278 (14). Additionally, one study reported the agreement between human raters and ChatGPT-3.5 with Fleiss’ kappa of 0.320, and between human raters and ChatGPT-4.0 with Fleiss’ kappa of 0.523 (13). Conversely, another study highlighted the poor acceptable reliability of ChatGPT (20), and one study reported a Cohen’s kappa of 0.341 for the agreement between ChatGPT and the reference standard (16). The most commonly identified limitation across the studies was the small sample size. **(Table 1)**

**Table 1.** Data extraction table.

| First author | Publication Year | Study Design | Type of medical cases triaged | Number of evaluated scenarios | Country | Large Language Model Used | Comparison Group | LLM's diagnostic performance | Inter-rater reliability | Main Findings | Limitations discussed in the included studies |
| --- | --- | --- | --- | --- | --- | --- | --- | --- | --- | --- | --- |
| Sinan Pasli et al. | 2024 | Observational study | Patients who presented to the ED | 758 | Turkey | ChatGPT 4.0 | Triage team and emergency medicine specialist | 77,77 % to 100 % sensitivity for triaging the patients | Near-perfect agreement with gold standard (Cohen's Kappa 0.899) for triaging | ChatGPT showed excellent predictive skills in triaging with high agreement with gold standard | Using local emergency rules, subjective general condition assessment |
| Rick Kye Gan et al. | 2023 | Cross-sectional study | Mass casualty incident cases | 15 | Malaysia | ChatGPT 3.5 and Google Bard | Medical students' performance from a previous study and Google Bard | ChatGPT: 66.67% over-triaged, Correctly-triaged 26.67%, 6.67% under-triaged; Google Bard: 60% Correctly-triaged, 40% over-triaged, no under-triage reported | Not reported | Google Bard was superior to ChatGPT in triage accuracy | Small sample size, only START triage protocol evaluated |
| Arian Zaboli et al. | 2024 | Observational study | Common cases in ED | 30 | Italy | ChatGPT 3.5 | Two triage nurses using Manchester Triage System (MTS) | Chat-GPT 3.5 - sensitivity (55.5%). Chat-GPT 3.5 - | Agreement in MTS code assignment between Comparison group and | This study showed Insufficient reliability and effectiveness of ChatGPT 3.5 to | Small sample size (30 vignettes), ChatGPT was not trained for ED triage. |
|  |  |  |  |  |  |  |  | specificity (57.1%) | Chat-GPT 3.5 was 0.278. (Cohen's Kappa) | replace triage nurses. |  |
| Jae Hyuk Kim et al. | 2024 | Inter-rater reliability study on virtual cases | Common cases in ED | 202 | Online (no specific location ) | ChatGPT 3.5 and ChatGPT 4.0 | 4 human raters (emergency staff) - experienced emergency medicine specialist | Not reported | Human raters with ChatGPT 3.5 (Fleiss' kappa=0.320) and Human raters with ChatGPT 4.0 (Fleiss' kappa=0.523) | ChatGPT have a Potential for ED triage but it's agreement with human raters is insufficient. | Use of virtual scenarios, not real patients |
| Jeffrey Michael Franc et al. | 2024 | Crossed gauge repeatability and reproducibility study | Common cases in ED | 61 | Online (no specific location ) | ChatGPT 3.5 | Emergency medicine specialists | ChatGPT 3.5: 47.5% Correctly-triaged, 13.7% under-triaged, and 38.7% over-triaged | Not reported | ChatGPT is not effective for triaging patients using the Canadian Triage and Acuity Scale, showing low repeatability and only 47% accuracy in its assessments. | One LLM evaluated, small number of prompts, prompts not optimized, no fine-tuning for triage |
| Marc Ayoub et al. | 2023 | Cross-sectional study | Common cases in ED | 9 | Online (no specific location ) | ChatGPT 3.5 | No direct comparison; Physicians scored responses | Average score 4.2/5 (SD 0.7) by 5 physicians for triage performance of ChatGPT 3.5 | Not reported | ChatGPT has the potential to augment clinical decision-making. | Small number of scenarios, clinician agreement difficult, need for further legal/accuracy evaluation |
| Max Ward et al. | 2024 | Comparative analysis - LLM vs. medical professionals | Cases needed Neurosurgical attention | 30 | USA | ChatGPT 3.5 and ChatGPT 4.0 | Neurosurgical attendings, residents, physician | ChatGPT 3.5: 92.59% Correctly-triaged; | Not reported | ChatGPT 4.0 performed at senior resident level and | Continuously changing nature of LLMs |
|  |  |  |  |  |  |  | assistants, subinterns | ChatGPT 4.0: 100% Correctly-triaged. |  | ChatGPT 3.5 performed near PGY-1 level for triaging the patients. |  |
| Stefan Bushuven et al. | 2023 | Cross-sectional study | Prehospital Basic Life Support and Paediatric Advanced Life Support Cases | 22 | Online (no specific location) | ChatGPT 3.5 and ChatGPT 4.0 | No human comparison | Not reported | Poor to acceptable reliability | ChatGPT/GPT-4 correctly identified 12 of 22 scenarios (54.5%) as emergencies of high urgency (Inconsistent emergency triage performance ). | No human comparison, selection bias, prototypical cases, regional variability |
| Hamish Fraser et al. | 2023 | Observational study | Patients who presented to the ED | 40 | USA | ChatGPT 3.5 and ChatGPT 4.0 | Ada symptom checker, WebMD symptom checker, ED physicians | ChatGPT 3.5: 59% Correctly-triaged, 41% under-triaged, and 0% over-triaged; ChatGPT 4.0: 76% Correctly-triaged, 22% under-triaged, and 3% over-triaged. | Not reported | ChatGPT 4.0 was better at triage than ChatGPT 3.5 but worse in diagnostic accuracy. | Data presentation differences, not simulating real patient queries, changes in versions |
| Riley J. Lyons et al. | 2023 | Cross-sectional study | Cases with common ophthalmic complaints | 44 | USA | ChatGPT 4.0 | Ophthalmology trainees, Bing Chat, WebMD Symptom Checker | ChatGPT 4.0: 98% Correctly-triaged. | Not reported | ChatGPT 4.0 showed high accuracy comparable to trainees for ophthalmology triaging the patient and There were no harmful statements generated with this LLM. | Probability of generating incorrect information from ChatGPT and propagating biases in triage |
| Rick Kye Gan et al. | 2023 | Cross-sectional study | Mass casualty incident cases | 15 | Online (no specific location ) | ChatGPT 3.5 | Medical students' triage performance | ChatGPT 3.5: 80% Correctly-triaged, 0% under-triaged, and 20% over-triaged | Not reported | ChatGPT 3.5 showed Higher triage performance compared to medical students after teaching | No explicit limitations identified |
| Georges Gebrael et al. | 2023 | Retrospective study | Metastatic prostate cancer patients | 56 | USA | ChatGPT 4.0 | Emergency medicine physicians | ChatGPT 4.0: 50% Correctly-triaged, 4% under-triaged, and 46% over-triaged | Not reported | ChatGPT 4.0 showed High sensitivity for admission of but lower specificity in discharges along with accurate diagnoses and treatment recommendations | Lower specificity in discharges, no association between ESI scores and outcomes |
| İbrahim Sarbay et al. | 2023 | Cross-sectional study | Common cases in ED | 50 | Turkey | ChatGPT 3.5 | Three emergency medicine specialists | ChatGPT 3.5: 60% Correctly-triaged, 18% under-triaged, and 22% over-triaged | Cohen's Kappa: 0.341 (fair agreement) | ChatGPT had better performance for high acuity cases | Small sample size, lack of generalizability, no validation on real data |
| Roya Zandi et al. | 2024 | In silico study | Common Ophthalmic Complaints | 80 | USA | ChatGPT 4.0 | Three ophthalmologists and Google Bard | ChatGPT 4.0: 85% Correctly-triaged | Not reported | GPT-4 outperformed Google Bard in triage and had lower potential for harm | Moderate sample size, couldn't fully assess hallucinatory responses or variability |

### Diagnostic Performance of ChatGPT for Triaging Patients

The following forest plots provide insights into the diagnostic performance of different versions of ChatGPT in triaging patients.

### ChatGPT 4.0 Performance

The pooled accuracy for ChatGPT 4.0 was 0.86 (95% CI: 0.64-0.98), with substantial heterogeneity among studies (I² = 93%). Individual study sensitivities ranged from 0.50 to 1.00. (**Figure** 2)

**Figure 2.**
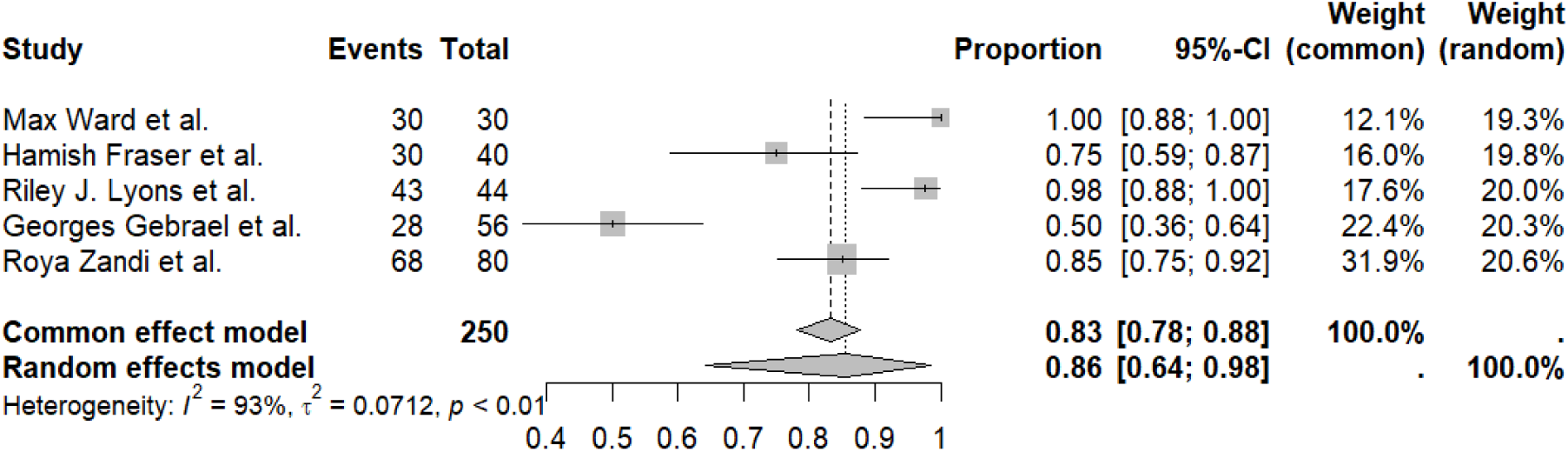
Forest Plot of ChatGPT 4.0 Performance.

### ChatGPT 3.5 Performance

The pooled accuracy for ChatGPT 3.5 was 0.63 (95% CI: 0.43-0.81), with significant heterogeneity (I² = 84%). Sensitivities in individual studies varied from 0.27 to 0.93. These results indicate that while ChatGPT 4.0 shows higher diagnostic performance for triaging ED patients compared to ChatGPT 3.5, there is considerable variability among studies. (**Figure** 3)

**Figure 3.**
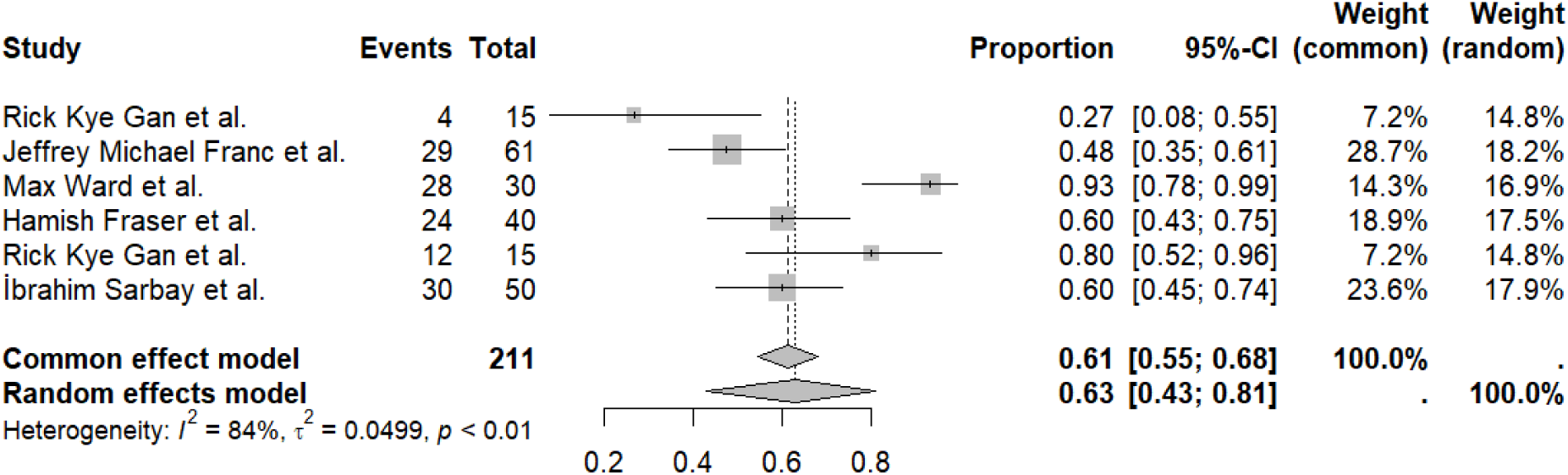
Forest Plot of ChatGPT 3.5 Performance.

### Sensitivity Analysis

Sensitivity analyses were conducted to evaluate the robustness of the pooled estimates. For ChatGPT 4.0, the sensitivity analysis (Figure 1S) demonstrated that omitting individual studies did not significantly alter the pooled accuracy, indicating stability in the results. For ChatGPT 3.5, the sensitivity analysis (Figure 2S) also showed consistent results, although the exclusion of some studies slightly affected the pooled estimates (19), reflecting moderate robustness. (**Figure 1S and Figure 2S**)

### Publication bias

Publication bias was assessed using funnel plots for both ChatGPT 4.0 and ChatGPT 3.5. The funnel plots (Figures 3S and 4S, respectively) showed some asymmetry, suggesting potential publication bias, particularly for ChatGPT 3.5. Due to the low number of studies, we couldn’t perform the Egger test for evaluating the funnel plot asymmetry. **(Figure 3S and Figure 4S**)

**Figure 4.**
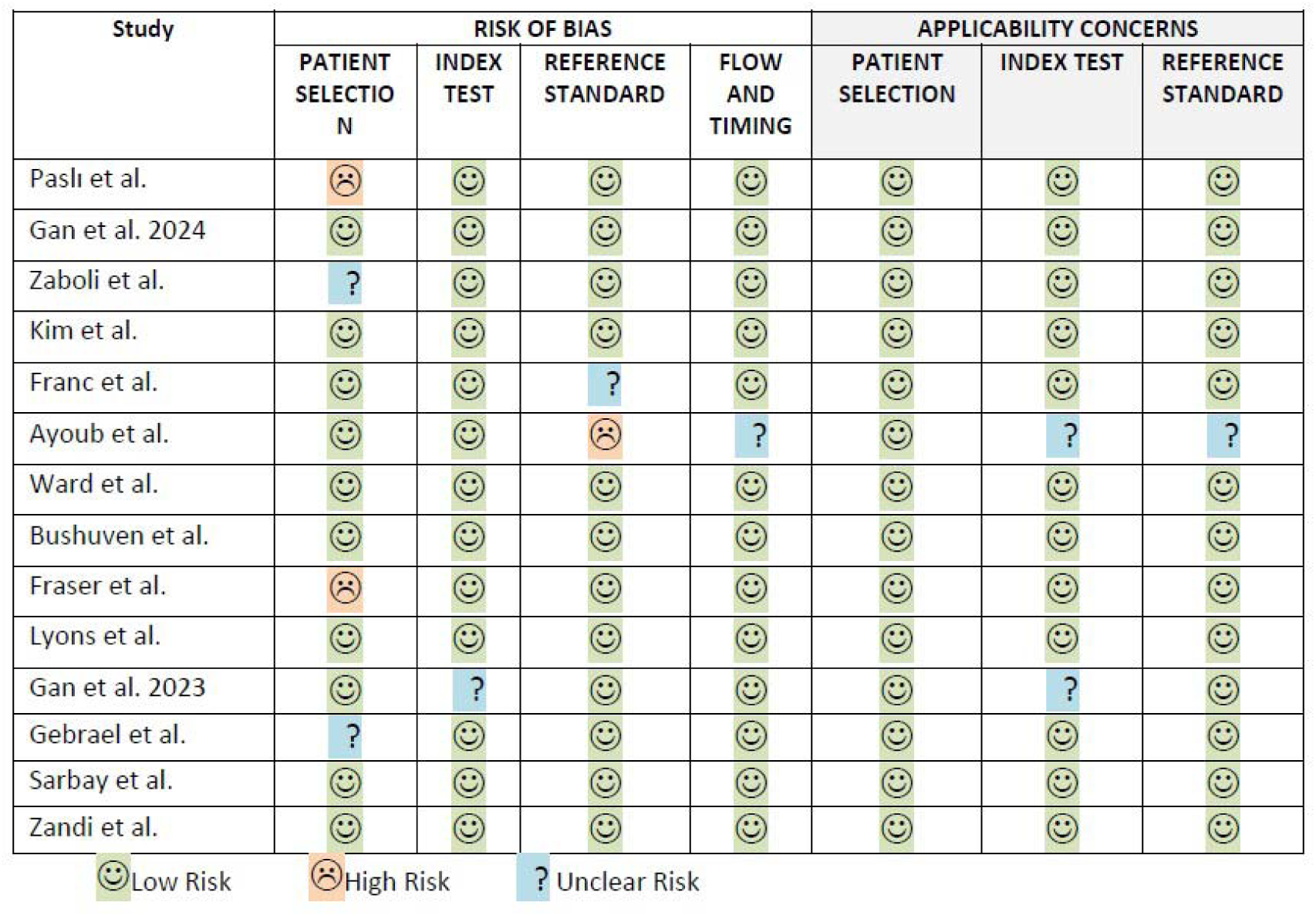
Quality Assessment using QUADAS-2.

### Quality Assessment

The quality assessment using the QUADAS-2 tool indicated varying levels of risk of bias and applicability concerns across the studies. Patient selection generally had a low risk of bias, though a few studies showed high or unclear risk. The index test was mostly low risk, indicating appropriate conduct of the tests. The reference standard was low risk in most studies. Flow and timing were also low risk in the majority of studies, indicating appropriate design execution. However, a few studies had high or unclear concerns regarding patient selection, index test, and reference standard applicability. (**Figure** 4)

## Discussion

The diagnostic performance of ChatGPT 4.0 was higher than ChatGPT 3.5, with pooled accuracies of 0.86 and 0.63, respectively. There was substantial heterogeneity among the studies for both versions. Sensitivity analyses indicated that the results were robust, though ChatGPT 3.5 showed some variability. Publication bias was suggested for both versions particularly ChatGPT 3.5 based on funnel plot asymmetry. The quality assessment using QUADAS-2 indicated varying levels of risk of bias and applicability concerns across the studies.

Our findings are consistent with those of Hirosawa et al. (24), who found a high diagnostic accuracy of 93.3% for ChatGPT 3.5 within differential-diagnosis lists, suggesting that AI chatbots can generate accurate diagnosis lists for common complaints. Rao et al. (25) also reported overall accuracy for ChatGPT at 71.7%, with the highest performance in final diagnosis and the lowest in initial differential diagnosis.

Mehnen et al. (26) noted that ChatGPT 4 requires more suggestions to solve rare diseases compared to common cases, aligning with our findings of variable diagnostic performance. Moreover, Williams et al. (27) found that GPT-3.5 achieved 84% accuracy in determining higher acuity patients.

The results indicate that ChatGPT, particularly version 4.0, has the potential to improve triage accuracy and reduce unsafe decisions in emergency settings. However, the variability among studies highlights the need for further evaluation and improvements. Fraser et al. (8) caution against unsupervised use of ChatGPT for triage without enhancements to accuracy and clinical validation. Gebrael et al. (23) suggest that ChatGPT can assist healthcare providers in improving patient triage, while Knebel et al. (28) found that although ChatGPT provides appropriate measures, there is a potential for harmful recommendations.

### Limitations

This meta-analysis provides a comprehensive evaluation of ChatGPT’s diagnostic performance in ED triage, incorporating a diverse range of studies and medical cases. However, the substantial heterogeneity and potential publication bias identified are limitations that must be considered. The varying levels of risk of bias and applicability concerns across studies further underscore the need for cautious interpretation of the results.

## Conclusion

This systematic review and meta-analysis assessed the diagnostic performance of ChatGPT 3.5 and ChatGPT 4.0 in emergency department (ED) triage. ChatGPT 4.0 demonstrated higher diagnostic accuracy (0.86) compared to ChatGPT 3.5 (0.63), but substantial heterogeneity and potential publication bias were noted. The variability in performance across different medical domains highlights the need for further evaluation and improvements.

## Acknowledgement

None

## Data Availability

Data of this study are available and will be provided if anyone needs them.

## Ethics approval and consent to participate

Not applicable

## Consent to publication

Not applicable

## Declaration of Generative AI and AI-assisted Technologies use in the Writing process

During the preparation of this work, the authors used ChatGPT 3.5 by OpenAI to improve paper readability. After using this service, the authors reviewed and edited the content as needed and took full responsibility for the publication’s content.

## Author contributions

All authors are accountable for all sections of the manuscript and declare that it is written originally and there is no data fabrication; data falsification including deceptive manipulation of images and plagiarism. Details of authors contributions are as follows:

(1) The conception and design of the study: Ramin Shahidi, Navid Kaboudi
(2) Acquisition of data: Fatemeh Fayazbakhsh, Salar Ghaderi, Mohammadreza Dehdashti, Maryam Vasaghi-Gharamaleki, Zahra Moradzadeh, Yasmin Mohtasham Kia
(3) Analysis and interpretation of data: Mohammad Shahir Eftekhar, Maryam Afshari, Fattaneh Khalaj
(4) Drafting the article: Ramin Shahidi, Niloufar Joharivarnoosfaderani, Zahra Mohammadi, Zahra Hasanabadi, Saeedeh Firouzbakht, Salar Ghaderi, Leila Haghani, Zahra Moradzadeh
(5) Revising it critically for important intellectual content: Ramin Shahidi, Navid Kaboudi, Saeedeh Firouzbakht, Leila Haghani
(6) Final approval of the version to be submitted: Ramin Shahidi, Navid Kaboudi, Leila Haghani Final version was read and approved by all author.

