## Supplementary material for "Diagnostic Performance of ChatGPT to Perform Emergency Department Triage: A Systematic Review and Meta-analysis"

Supplementary materials

1. Keywords
2. Search Queries
3. Figure 1S
4. Figure 2S
5. Figure 3S
6. Figure 4S

“LLM Keywords”

MESH Terms:

.

Emtree Terms:

large language model

ChatGPT

SYNONYMS:

Large language model

Bard

Gemini

Chat-GPT

ChatGPT

“Triage Keywords”

MESH Terms:

Triage

Emtree Terms:

patient triage

SYNONYMS:

Triage*

triaging

Pubmed Search Query

Time of search : 19 March 2024
Results : 39

(Large language model[tiab] OR Bard[tiab] OR Gemini[tiab] OR Chat-GPT[tiab] OR ChatGPT[tiab]) AND ("Triage"[mesh] OR Triage*[tiab] OR triaging[tiab])

Embase Search Query

Time of search : 19 March 2024
Results : 55

(‘large language model’/exp OR ‘ChatGPT’/exp OR ‘Large language model’:ab,ti OR ‘Bard’:ab,ti OR ‘Gemini’:ab,ti OR ‘Chat-GPT’:ab,ti OR ‘ChatGPT’:ab,ti) AND (‘patient triage’/exp OR ‘Triage*’:ab,ti OR ‘triaging’:ab,ti)

Scopus Search Query

Time of search : 19 March 2024
Results : 38

(TITLE-ABS (“Large language model” OR “Bard” OR “Gemini” OR “Chat-GPT” OR “ChatGPT”)) AND (TITLE-ABS (“Triage*” OR “triaging”))

Web of science Search Query

Time of search : 19 March 2024
Results : 30

(TS= (“Large language model” OR “Bard” OR “Gemini” OR “Chat-GPT” OR “ChatGPT”)) AND (TS= (“Triage*” OR “triaging”))

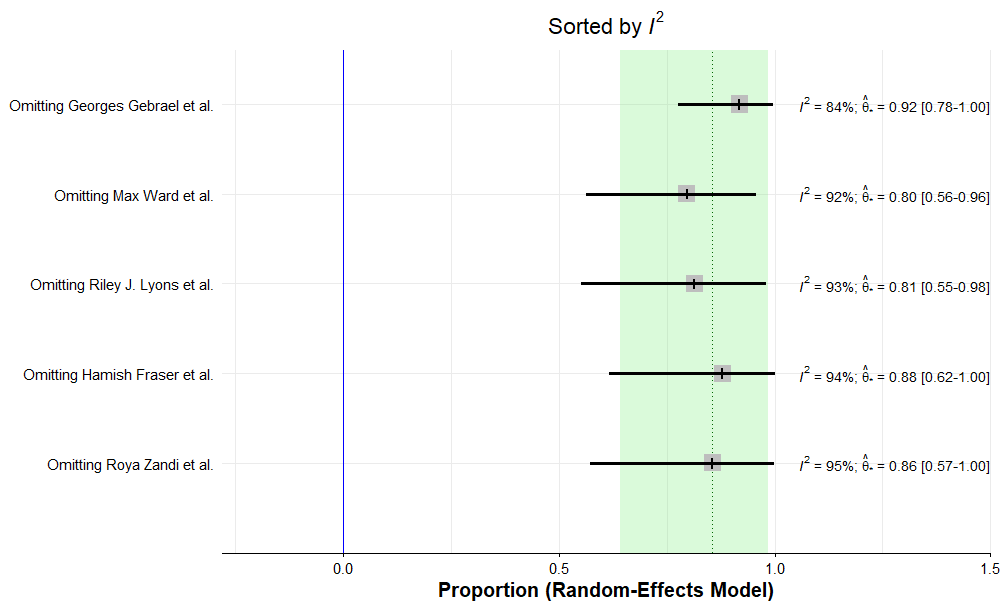

**Figure 1S. Sensitivity analysis of ChatGPT 4.0 pooled accuracy**

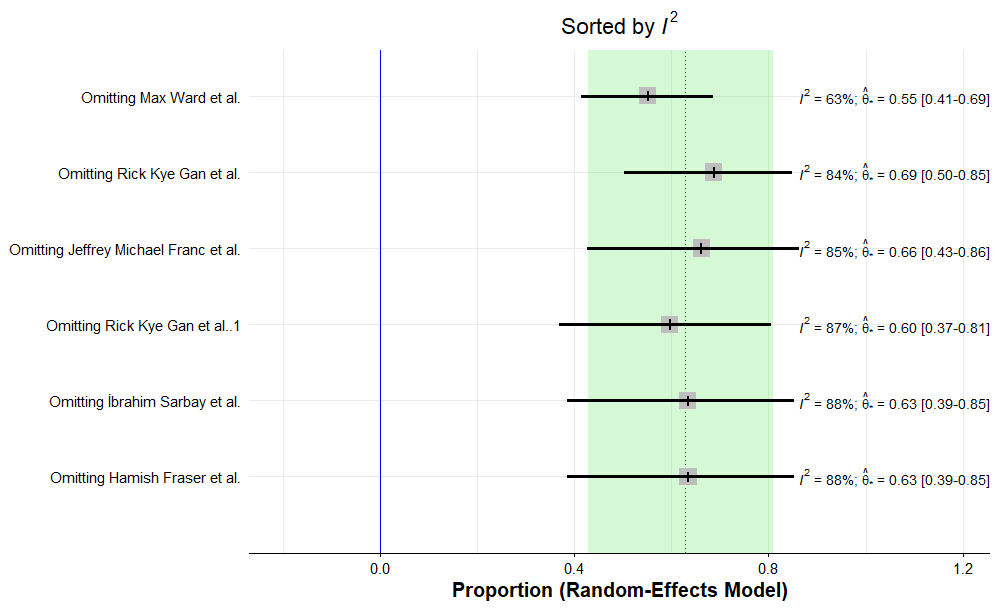

**Figure 2S. Sensitivity analysis of ChatGPT 3.5 pooled accuracy**

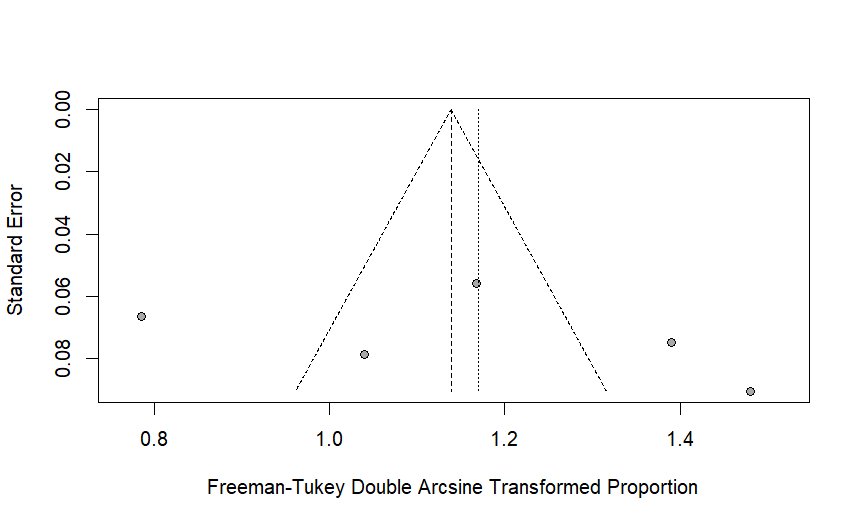

**Figure 3S. Funnel plot of ChatGPT 4.0**

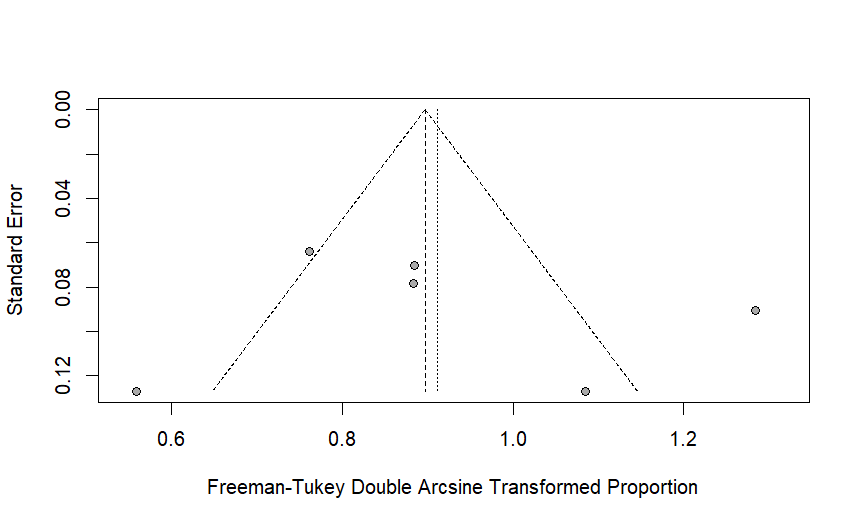

**Figure 4S. Funnel plot of ChatGPT 3.5**
